# A systematic review of clinical data and reporting quality in NMDAR-antibody encephalitis and pregnancy

**DOI:** 10.1101/2024.11.28.24317822

**Authors:** Scarlett L Harris, Sophie NM Binks, Donal Skelly, Hanine Fourie, Phoebe Cherrington-Walker, Tomasz Bajorek, Sarosh R Irani, M Isabel Leite, Adam E Handel, Adam Al-Diwani

**Affiliations:** Medical Sciences Division, University of Oxford, Oxford, UK; Faculty of Medicine, Imperial College London, London, UK; Oxford Autoimmune Neurology Group, Nuffield Department of Clinical Neurosciences, University of Oxford, Oxford, UK; Department of Neurology, Oxford University Hospitals NHS Foundation Trust, Oxford, UK; Oxford Centre of Neuroinflammation, Nuffield Department of Clinical Neurosciences, University of Oxford, Oxford, UK; Department of Obstetrics and Gynaecology, Oxford University Hospitals NHS Foundation Trust, Oxford, UK; Departments of Neurology and Neuroscience, Mayo Clinic, Jacksonville, Florida, USA; Department of Psychiatry, University of Oxford, Oxford, UK

## Abstract

**Background:** NMDAR antibody encephalitis (NMDAR-Ab-E) can have an onset during, after, or prior to a pregnancy. In animal models, transplacental NMDAR-IgG transfer can affect neurodevelopment. In contrast, clinical reports of mothers affected by NMDAR-Ab-E typically are reassuring. We systematically reviewed maternal, infant, and childhood clinical data pertaining to NMDAR-Ab-E with an onset before, during, or after pregnancy and compared this to our single autoimmune neurology centre experience.

**Methods:** After pre-registration on PROSPERO (CRD42023408447), we searched PubMed and Scopus for NMDAR-Ab-E case reports/series with an onset before, during, or after pregnancy (last search 19/10/2023). We extracted maternal, neonatal, and childhood outcomes using an idealised checklist to derive summary statistics.

**Results:** After quality control we identified 66 pregnancies in 61 women from 48 reports or series. 72% of women recovered with minimal or no neurological deficits, comparable to non-pregnancy associated NMDAR-Ab-E. Likewise, 80% of pregnancies resulted in livebirths with a single neonatal death reported. Data on neonatal outcome measures were frequently unreported and childhood follow-up in only 60%. Our centre’s experience is consistent: 3/4 mothers recovered with no functional deficits and 7/8 children without evidence of compromise at median of two years follow-up.

**Conclusions:** Current evidence does not overall suggest unfavourable maternal, fetal, or childhood outcomes after NMDAR-Ab-E. However, the available sample is small, predominantly single case reports with modest follow-up, lacks standardisation, and data are often incomplete. Future approaches should address these caveats; developing multi-centre collaboration towards an international registry.

**Key messages:** *What is already known on this topic:* Some animals models of NMDAR-IgG transplacental transfer show adverse effects on brain development. However, caveats include species differences and potentially non-physiological exposures. Moreover, although some case reports identify adverse maternal and fetal outcomes, previous systematic reviews and single centre summaries of clinical data have been more reassuring.

*What this study adds:* We update and expand upon previous systematic reviews by including cases of NMDAR-Ab-E in the postpartum period and cases of pregnancy after recovery, as well as reporting the experiences of our autoimmune neurology centre. Additionally, we also focus on childhood outcomes and have contacted authors of published case reports for further follow-up. These data show generally good outcomes for mothers and children but reporting is patchy and not standardised.

*How this study might affect research, practice or policy:* To overcome these shortcomings in reporting we recommend collaboration amongst the autoimmune neurology clinical-research community to consolidate experience. This could include establishing an international registry to foster reporting standardisation and improve understanding of interactions between the illness, pregnancy, and potential effects on neonatal and childhood outcomes.

## Introduction

N-methyl-D-aspartate receptor antibody encephalitis (NMDAR-Ab-E) is an autoimmune neurological disorder predominantly affecting women of reproductive age (1,2). Mediated by IgG autoantibodies against the NR1 (GluN1) subunit of the NMDA receptor (NMDAR-IgG), NMDAR-Ab-E presents with combinations of acute psychiatric disturbance, movement disorders, seizures, dysautonomia, hypoventilation, and altered level of consciousness. Increasingly, this condition has been identified during pregnancy or in the postpartum period (3). Additionally, many who recover from the illness have yet to start or complete their family. They and their clinicians require clarity on potential risks for both mother and baby.

NMDAR-IgGs are typically of the IgG1 subclass. IgG1 autoantibodies can cross the placenta and induce congenital disease including in the nervous system. For example, in myaesthenia gravis autoantibodies against fetal acetylcholine receptor isoforms can cause fetal acetylcholine receptor antibody-related disorders, a spectrum of disorders ranging from milder myopathic presentations to arthrogryposis multiplex congenita (4). In these cases, immunomodulation, particularly early in pregnancy, has been shown to improve survival and reduce complications for the developing foetus. In an era of autoantibodies against central nervous system targets a similar question has been posed of fetal brain development. In animal models, CASPR2 and NMDAR-autoantibodies have been shown potentially to affect neurodevelopment (5,6). Furthermore, NMDAR-IgG seropositivity often persists despite clinical remission (7,8) and so syncytiotrophoblastic neonatal FcRn receptors could mediate transfer of the dominant IgG1 sub-class autoantibodies (9,10). Nonetheless, real world clinical outcomes have been more reassuring. For example, a previous systematic review found 10/13 livebirths with 8/10 healthy neonates (3) and an experienced autoimmune neurology centre reported 10/11 neonates healthy at birth (11). Moreover, in cases where there has been proven NMDAR-IgG transfer with sub-optimal neonatal outcomes, potential confounders have included maternal condition, medication, and placental factors (12,13).

Here, we aimed to assess maternal, fetal/neonatal, and childhood outcomes with a focus on reporting quality to inform recommendations on future standards. We deployed an idealised checklist of features in pregnancy and developmental features to systematically review literature-reported cases and compare with experience from our own autoimmune neurology centre.

## Methods

We pre-registered the study protocol with NIHR PROSPERO on 17/3/2023 (CRD42023408447) and followed Preferred Reporting Items for Systematic reviews and Meta-Analysis (PRISMA) guidance.

### Search strategy

We searched two databases (PubMed and Scopus) without language or date restriction using the search terms *(“anti-NMDA receptor” OR “anti-NMDAR” OR “anti-N-methyl-D-aspartate receptor encephalitis” OR “NMDAR-antibody encephalitis” OR “NMDAR-Ab-E” OR “NMDAR encephalitis” OR NMDARe) AND (pregnancy OR postpartum OR post-partum OR puerperal OR puerperium OR foetus OR fetus OR gestation OR birth OR neonate OR infant OR child OR perinatal).* We screened the reference lists of included papers for additional publications. The search was repeated twice to identify any papers published prior to the final analysis.

### Eligibility criteria

We included case reports and series which reported on patients with an onset of NMDAR-Ab-E *before* (‘non-pregnancy-associated’), or *during* or *after* pregnancy (‘pregnancy-associated’), as well as reports of children born to these patients. We planned to restrict to cases that strictly met the 2016 consensus criteria for definite anti-NMDAR encephalitis (14). However, our initial search yielded seven cases, including four published prior to these criteria, which did not fully meet definite classification due to not measuring CSF NMDAR-IgG. Yet being highly typical for the illness they met probable criteria, and given the modest sample size and valuable clinical information therein, we chose to include these cases.

We initially defined postpartum onset as within 42 days as per WHO (15). However, only two of eight postpartum cases occurred within this period. Further aiming to maximise the inclusion of clinically-relevant information, we extended the postpartum definition to include cases where the presenting disorder was classified as postpartum in onset, which here was a maximum of 11 months postpartum.

### Outcome measures

The full template for data collection including all extracted outcomes is provided in Supplementary Table 1. As primary outcomes we aimed to ascertain maternal morbidity, mortality, and functional status, pregnancy complications, and morbidity, mortality, and functional status in neonates (baby <28 days old), and where available, later childhood developmental progress. We defined preterm birth as before 37 weeks and low birth weight at term as <2.5 kg as per WHO (16). We defined postpartum haemorrhage as blood loss >500ml as per RCOG (17). We defined normal CSF parameters as protein concentration 15-40 mg/dL and white cell count 0-5/mm^3^ (18). As secondary outcomes, we noted whether NMDAR-IgG autoantibodies were reported in cord or neonatal blood samples alongside maternal serology.

### Data extraction

We removed duplicate papers to produce a final list of abstracts for screening. Two authors (SH, AAD) independently compared a representative sample (n=21) of the abstracts and reached consensus on inclusion with full agreement. The remaining abstracts were screened yielding 48 papers eligible for inclusion. SH performed the extraction which was then independently cross-checked (AAD, DS, and HF). Any differences were resolved by discussion. Where data was insufficient, we contacted the report authors to supplement the available published data.

### Quality Assessment

Studies were assessed for quality using the tool for evaluating the methodological quality of case reports and case series (19). We made project-specific modifications to prioritise whether there was sufficient information to: 1) confirm the diagnosis of NMDAR-Ab-E and 2) allow basic evaluation of neonatal outcomes (Supplementary Table 1). Maternal outcome data was not used to determine inclusion as we did not wish to exclude records of children born secondary to pregnancies complicated by NMDAR-Ab-E which may not report maternal outcome. Those providing information on childhood outcome and with follow-up of at least one year were considered good quality.

### Local case series

To contextualise the global experience from the systematic review we reported cases from our autoimmune neurology service who satisfied the same eligibility criteria. All are participants in the Immune Factors in Neurological Disease research study (REC 16/YH/0013) and gave informed consent in accordance with the Declaration of Helsinki. If an assenting participant lacked capacity to consent for themselves then there was a next of kin declaration. Additional publication-specific consent was obtained for de-identified detailed individual participant data including offspring.

### Data analysis

Data were tabulated with Excel version 16.83 (Microsoft). Statistical analyses and visualisation were conducted with Prism version 10.2.1 (GraphPad). Fisher’s exact test was used to compare between pregnancy groups and the binomial or Chi-square tests to compare observed to expected results. Statistical significance was inferred where P <0.05.

## Results

### Identification of records

Our initial search identified 1587 records (733 PubMed; 854 Scopus, Fig. 1). Later repeated searches identified an additional 107 records (49 PubMed, 58 Scopus). 638 duplicate records were removed leaving 1056 for abstract screening. This was then refined to 60 eligible records including two identified through abstract screening (Supplementary Table 2). Two systematic reviews and two papers with insufficient data were removed.

**Figure 1:**
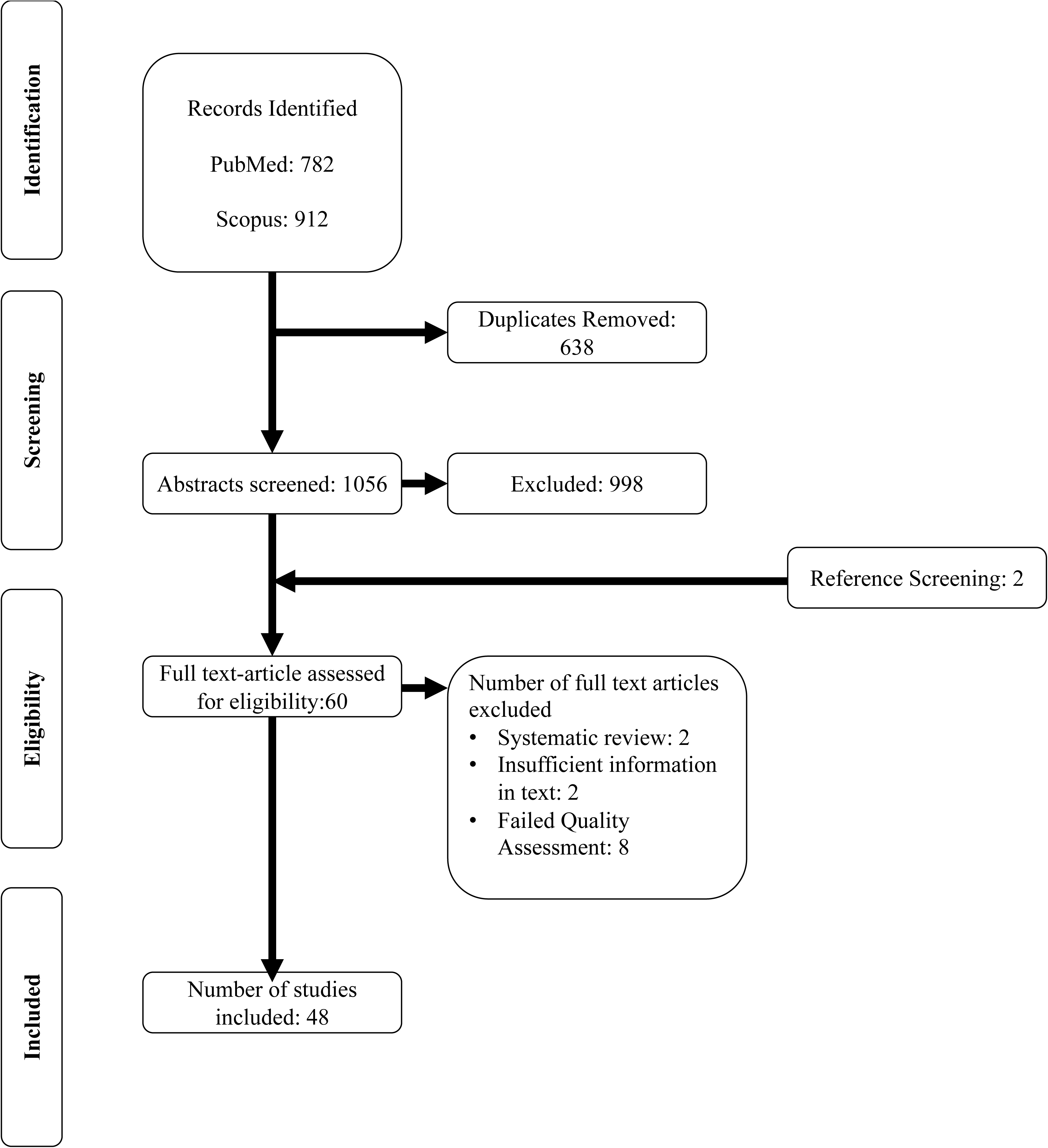
PRISMA flow diagram. The combined findings of the initial search conducted on the 21^st^ of March 2023 and subsequent searches on the 17^th^ of July and 19^th^ of October 2023 are shown. After screening, removal of duplicates, and quality control, 48 studies were identified as eligible for inclusion.

We then quality-assessed the remaining 56 records, finding 48 of sufficient quality for inclusion (n=8 inadequate, n=33 adequate, n=15 good; Fig. 1). There were 53 NMDAR-Ab-E cases associated with pregnancy including 45 with an onset *during* pregnancy involving 43 individual women with two cases of relapse *during* subsequent pregnancies. There were eight cases *after* pregnancy. There were 13 non-pregnancy-associated cases occurring *before* a pregnancy including two women who had NMDAR-Ab-E during a previous pregnancy, and one woman who had two pregnancies after recovery.

### NMDAR-Ab-E description and treatment

The average age of illness onset was consistent across the groups (Fig. 2). The overall ovarian teratoma rate in the pregnancy-associated cases was 43% (23/53) comparable to the literature-reported rate (20). This comprised 17/45 (38%) *during* and 6/8 (75%) *after* (non-significant; P = 0.065, Fisher’s exact test).

**Figure 2:**
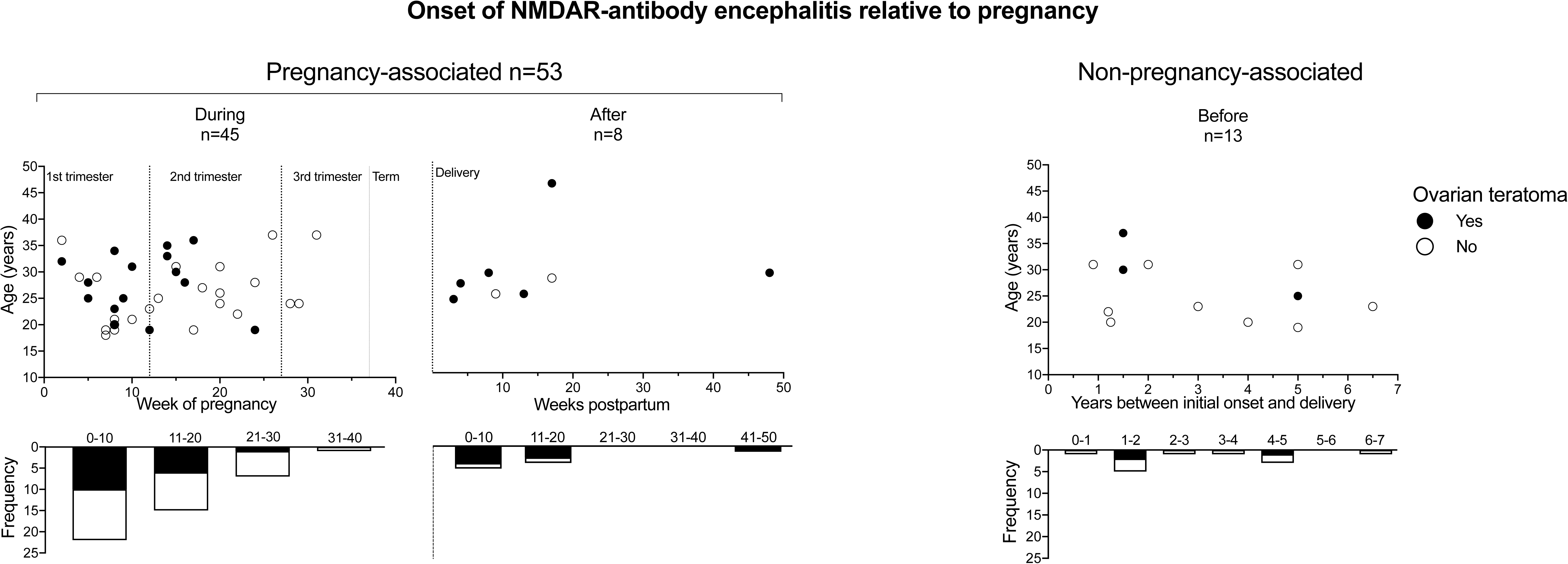
Onset of NMDAR-antibody encephalitis relative to maternal age and pregnancy. Cases of NMDAR-antibody encephalitis are plotted according to stage of pregnancy and maternal age. They are split into pregnancy associated (illness onset *during* or *after* pregnancy; left) and non-pregnancy associated (illness onset *before* pregnancy; right). For *during* cases, 41 of the 45 cases are represented since four cases did not give a specific onset time in gestational weeks but were all within the first trimester and two were associated with a teratoma. One teratoma-associated *before* case is not shown since the time interval was not clearly stated.

The majority of cases that occurred *during* pregnancy were early with only four in the third trimester (9% (4/45), P = 0.001, Chi-Square test). The overall clinical profile of the cases associated with pregnancy versus non-pregnancy associated cases differed in autonomic dysfunction (40% vs 10%, P = 0.145, Fisher’s exact test), reduced consciousness (70% vs 10%, P = 0.0006, Fisher’s exact test), and hypoventilation (49% vs 10%, P = 0.034, Fisher’s exact test) (Supplementary Fig. 1A). The rates of these features in the pregnancy-associated group is broadly in keeping with a recent large meta-analysis of 1550 predominantly female patients, where reduced level of consciousness was reported in 55% and autonomic dysfunction or central hypoventilation in 43% (20). Investigation and treatment profiles were also broadly similar across case types (Supplementary Fig. 1B-D).

### Pregnancy outcomes

For cases with an illness onset *during* pregnancy, most resulted in live births (33/45, 73%) (Fig. 3A). However, 18/33 (55%) were preterm (median gestational age 33 weeks, range 27-36) of which most were iatrogenic, i.e. either induced or involved a caesarean section (14/18, 78%; Fig. 3A). Fourteen women had a livebirth at term (14/33, 42%) and 29% (4/14) of these babies were born by caesarean section. We plotted available birth weights against week of delivery and these clustered at or below the normative median with 1/12 >97^th^ centile and 2/12 <3^rd^ centile (Fig. 3C). One paper reported a birthweight of 408g at 33 weeks which is close to the limit of viability and appeared implausibly low (21). Attempts to contact the authors to clarify were unsuccessful therefore we elected to exclude this data point from this analysis (retained and compared for reference in Supplementary Fig. 2).

**Figure 3:**
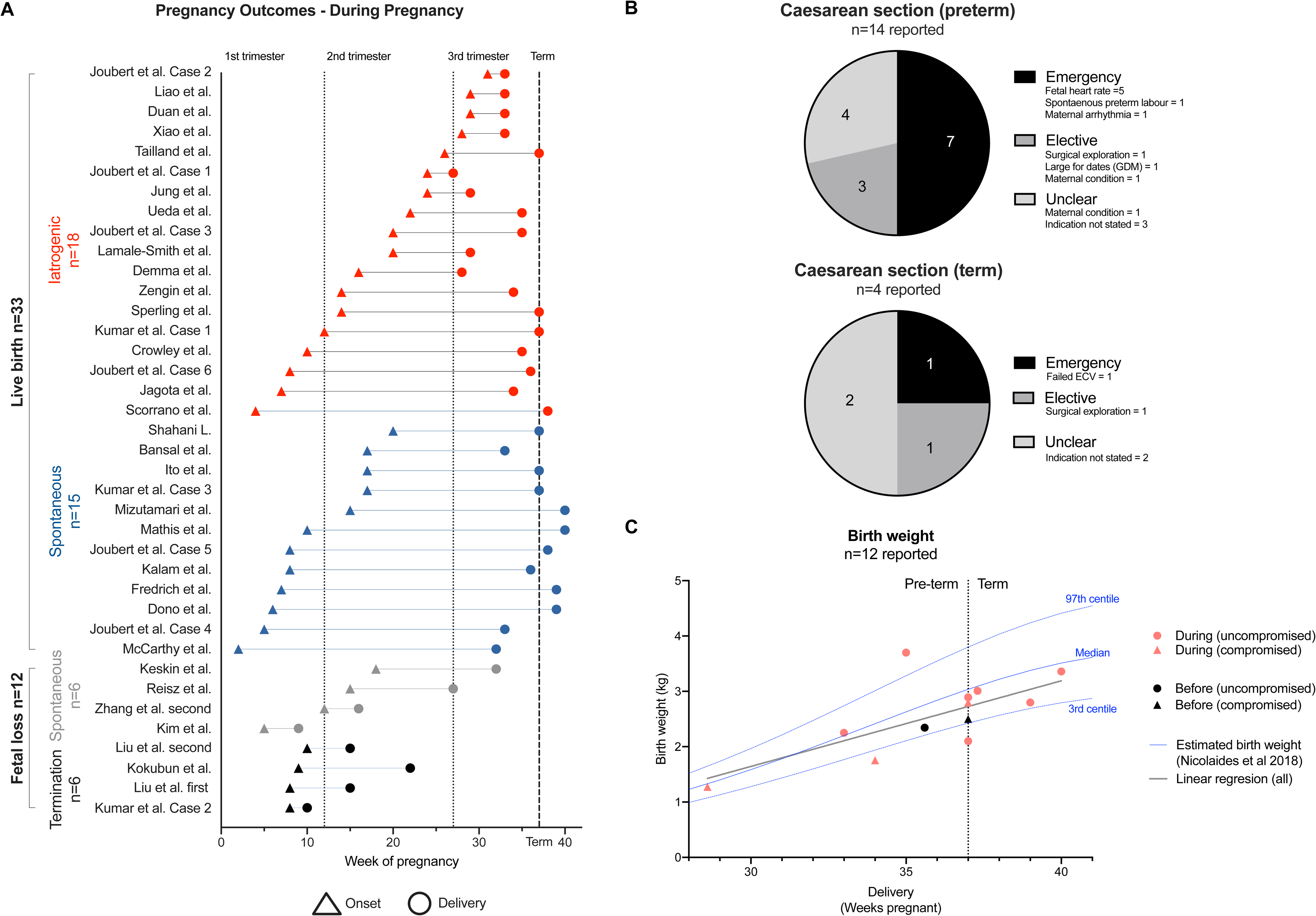
Pregnancy outcomes. A – Cases of NMDAR-antibody encephalitis during pregnancy are shown according to illness onset in relation to gestational time and pregnancy outcome (gestational week at presentation = triangles; end of pregnancy = circle; termination = black; spontaneous fetal loss = grey; spontaneous livebirth = blue; iatrogenic livebirth = red). Vertical dotted lines represent the end of the first and second trimesters and the dashed line at week 37 demarcates term. Four first trimester cases of fetal loss (two termination and two miscarriage) are not shown as specific timings were unavailable. Three spontaneous livebirths are not shown since specific delivery time was not described (36–38). B – In cases with an illness onset during pregnancy, pie charts summarise the proportion and absolute number of indications for caesarean section (preterm deliveries, top; term deliveries, bottom). C – Twelve available birthweights are plotted by gestational week (onset *during* pregnancy pink, onset *before* pregnancy black; uncompromised=circle; compromised=triangle) with a line of best fit (grey) in the context of normative birth weight ranges (blue lines; as per Nicolaides et al Ultrasound Obstet Gynecol 2018; 52: 44–51).

Six pregnancies ended spontaneously (four miscarriages and two stillbirths) and another six were terminated (Supplementary Table 3). In the non-pregnancy-associated *before* group there was one termination and the rest were livebirths (12/13, 92%) with two (17%) caesarean sections. All the cases with illness onset *after* pregnancy were livebirths with no sections reported. Generally, antenatal and delivery outcomes were rarely reported (Supplementary Fig. 3).

### Maternal outcomes

Most women recovered fully or with minimal neurological deficit (*before* 8/10, 80%; *during* 31/43, 72%; *after* 5/8, 63%). However, follow-up duration was modest, with only 12/40 (30%) of the *during* cases reporting maternal follow-up for more than a year. Across all the cases there were four maternal deaths (4/61, 6.6%; Fig. 4A – top and Supplementary Table 4). These were predominantly secondary to sepsis, a relatively common cause of maternal death that accounted for 10% of maternal deaths in the UK between 2019-2021 (22).

**Figure 4:**
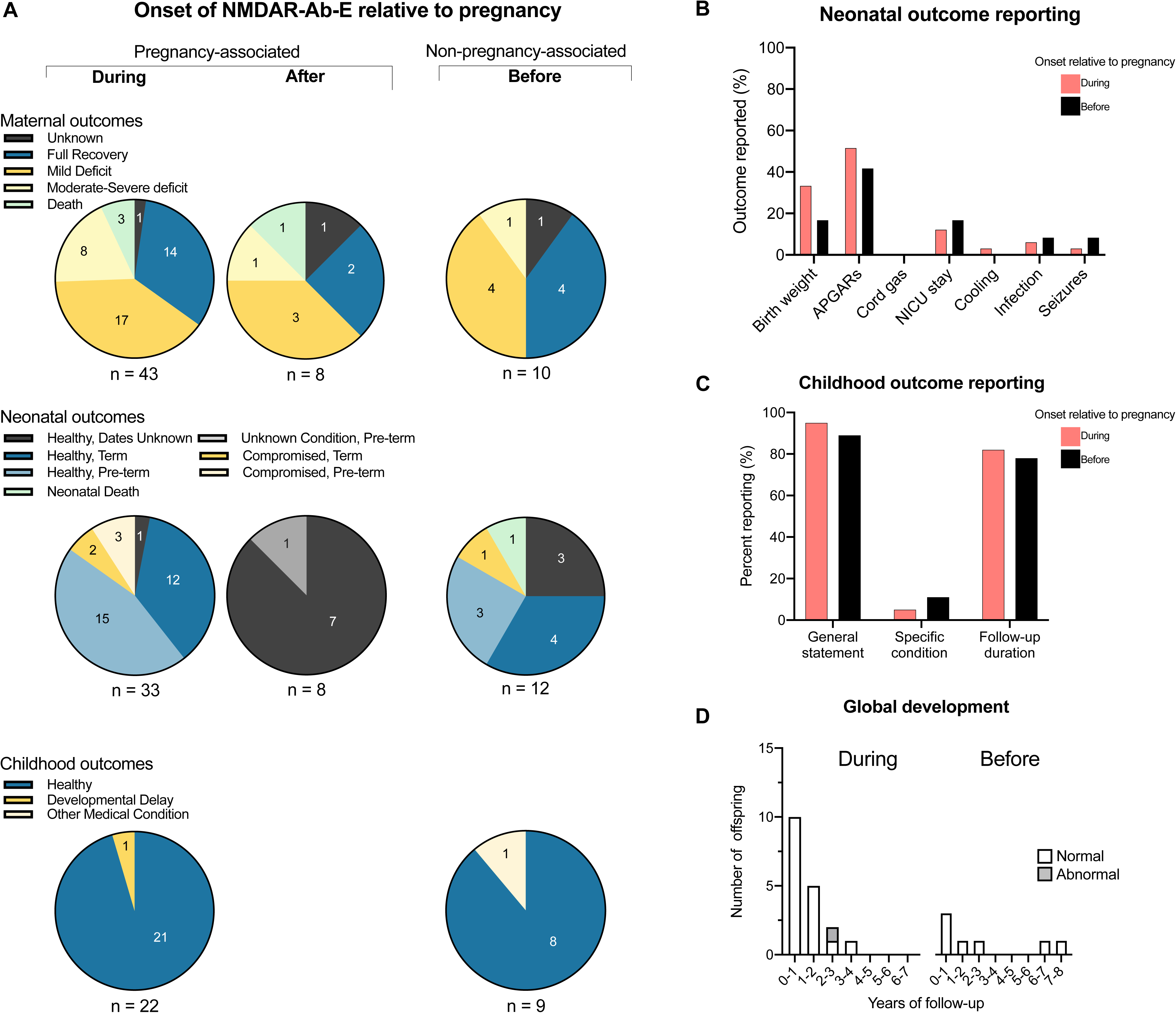
Maternal, neonatal and childhood outcomes. A – Maternal, neonatal and childhood outcomes are shown divided by onset of NMDAR-antibody encephalitis in relation to pregnancy. For maternal outcomes, cases where deficit was reported as “minimal” or modified Rankin score (mRS)=1-2 are shown as mild deficit. Cases are defined as moderate-severe deficit for mRS 3-5 or clear functional disability described. For neonatal outcomes, infants are specified as compromised if this was specifically stated or as indicated by Apgar scores. B – Positively reported neonatal outcomes are plotted as percentages for pregnancies with an onset of NMDAR-Ab-E occurring *during* (pink) or *before* (black). C – Reported childhood outcomes are plotted as percentages for pregnancies with an onset of NMDAR-Ab-E occurring *during* (pink) or *before* (black). General statement refers to a descriptor of adequate progress such as “healthy”, “met all developmental milestones”, “developing normally”. Also plotted are statements of a specific condition and whether duration of follow-up was stated. D – Where available, specific duration of childhood follow-up of global development is plotted as a bar chart for pregnancies with an onset of NMDAR-Ab-E occurring *during* (left) or *before* (right). Offspring in whom an abnormality was reported are shaded grey. Abbreviations: APGAR=Appearance, Pulse, Grimace, Activity and Respiration, mRS=modified Rankin score, NMDAR-Ab-E=NMDAR-antibody encephalitis, NICU= neonatal intensive care unit.

### Neonatal outcomes

Overall, while there was little evidence for poor neonatal outcomes (Fig. 4A – middle), reporting of specific neonatal outcomes was generally sparse (Fig. 4B). This was especially true for *after* cases where there was only one case of preterm birth reported, secondary to placental abruption (Fig. 4A – middle). In three of these cases no specific statement on neonatal health was given, but we could infer the neonate was alive because the mother was breastfeeding, or the delivery was described as normal.

Overall, 7/53 (five *during* cases and two *before* cases; 13%) neonates were identified as compromised (Supplementary Table 5). Of these seven, five were tested for NMDAR-IgG, of which four (80%) were positive. Five healthy neonates were tested with only one positive (Fig. 5A-B). There was one death reported of an already compromised neonate. The mother had previously recovered from NMDAR-Ab-E, although the interval to the pregnancy was relatively short with the illness onset preceding delivery by 18 months (13). Whilst there were concerns regarding an encephalitis relapse, the patient was unaware of her pregnancy and presented with unmodified hypertension and deranged liver function consistent with pre-eclampsia, indicative of a likely confounding aetiology.

**Figure 5:**
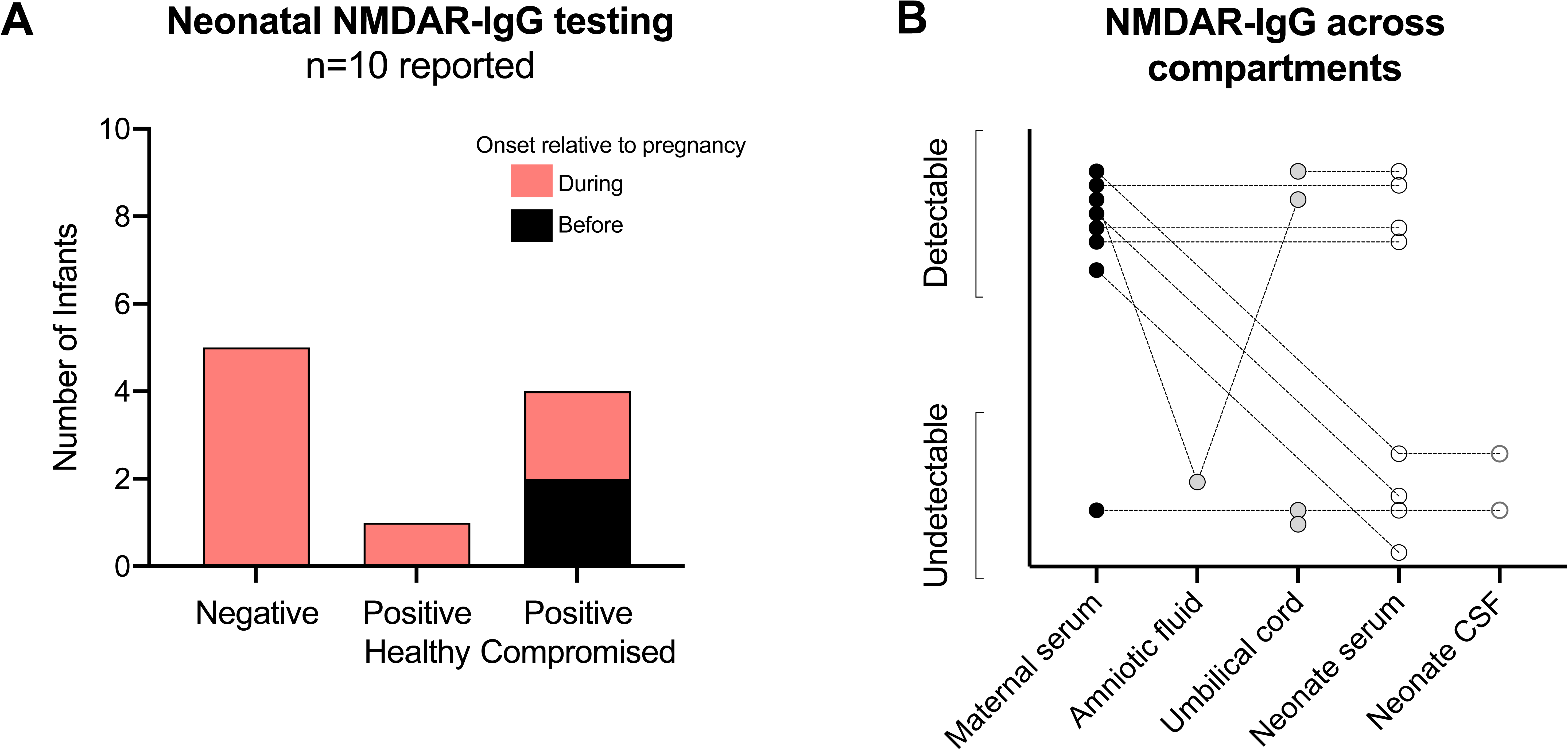
Neonatal NMDAR-autoantibody testing. A – The absolute number of infants tested for NMDAR IgG autoantibodies are plotted according to the test result. Positive results are divided according to whether the infant was clinically healthy or compromised at the time of testing. The bars are sub-divided according to whether NMDAR-Ab-E occurred *during* (pink) or *before* (black) the associated pregnancy. B – Results of NMDAR-IgG assays are plotted according to corresponding bio-fluid tested. Dotted lines note samples connected within a maternal-neonatal pair. Abbreviations: CSF=cerebrospinal fluid, NMDAR-IgG=Immunoglobulin G autoantibody against N-Methyl D-Aspartate receptor

### Childhood outcomes

Of the 33 cases in which illness began during pregnancy ending with a live birth, childhood outcomes were provided in 22 (Fig. 4A – bottom). The level of detail was largely restricted to a general statement in most cases that the children were healthy and/or meeting developmental milestones (Fig. 4C). However, the duration of follow-up and therefore opportunity to identify more complex neuro-developmental outcomes was limited. For cases with an illness onset during pregnancy 8/22 (36%) were followed-up longer than a year, whilst this figure was 44% for those born to mothers who had recovered from NMDAR-Ab-E prior to pregnancy (4 of 9 cases providing childhood follow-up, Fig. 4D). There was no childhood data provided for cases with an illness onset after pregnancy.

Amongst *during* cases, one child had global developmental delay and in this case the mother’s illness was severe and she died secondary to infection (12). The baby’s serum was positive for NMDAR-IgG at birth but negative by one year. This child was identified as compromised in the neonatal period, but the other three compromised neonates for whom data was available, went on to develop normally. For the *before* cases only one child had reported medical diagnoses, which were torticollis and strabismus (23). This child was not identified as compromised at birth and the one surviving compromised child was described as developmentally normal. Maternal, neonatal, and child outcomes are summarised in Supplementary Fig. 4.

Given the number of potential childhood outcomes left unreported and limited length of reported follow-up we contacted authors to ascertain if further follow-up was available. 7 of 27 authors contacted responded, of which three were able to provide further follow-up. No new diagnoses were made to alter the existing reported literature.

### Oxford autoimmune neurology experience

In addition to a group summary (Table 1), where possible we obtained specific consent to report de-identified individual participant data according to our checklist (Supplementary Table 6). We have not encountered any patients with a postpartum onset of the illness but two cases with an onset *during* pregnancy, a relapse and first illness. Both neonates were born premature but live (one spontaneous delivery and one emergency C-section secondary to non-reassuring fetal heartbeat). Both were admitted to the special care baby unit and have developed along normal trajectories with a median follow-up of 1.5 years.

**Table 1.** Group summary of single centre experience of NMDAR-Ab-E and pregnancy with maternal, neonatal, and childhood outcomes.

| NMDAR-Ab-E onset relative to pregnancy | Number of patients, n | Number of pregnancies, n | Trimester at NMDAR-Ab-E onset | Miscarriage, n | Live births, n | Birth timing years post illness onset, median (range) | Preterm birth, n | Term birth, n | SCBU admission, n | Years maternal follow-up since episode, median (range) | Maternal outcome mRS 0-1, n | Years child follow-up, median (range) | Child outcome within normal limits, n |
| --- | --- | --- | --- | --- | --- | --- | --- | --- | --- | --- | --- | --- | --- |
| Before | 3 | 7 | N/A | 1 | 6 | 3.5 (1.5-6) | 0 | 6 | 1* | 7 (2-12) | 3 | 1.75 (0.5-5) | 5 |
| During | 2 | 2 | 1<sup>st</sup> and 2<sup>nd</sup> | 0 | 2 | N/A | 2 | 0 | 2 | 1.5 (1-2) | 1 | 1.5 (1-2) | 2 |
| After | 0 | 0 | N/A | N/A | N/A | N/A | N/A | N/A | N/A | N/A | N/A | N/A | N/A |

\*Phototherapy for neonatal jaundice in context of PPRM
Abbreviations: mRS=modified Rankin score, NMDAR-Ab-E=NMDAR-antibody encephalitis, n=number, N/A = not applicable, SCBU=special care baby unit

Beyond this, most of our experience is non-pregnancy-associated NMDAR-Ab-E, occurring and resolving *before* pregnancy (seven pregnancies from three mothers). All mothers had recovered without residual deficit, and six pregnancies resulted in term livebirths, with one miscarriage. Four of the six neonates were born in good condition. One had raised respiratory rate at birth and was treated for possible sepsis. Heel prick blood from this neonate was positive for NMDAR-IgG, but there have been no developmental concerns with five years of follow-up. The other was treated for sepsis and jaundice in the context of preterm premature rupture of membranes (PPROM). Here, development has been largely as expected but an assessment for potential neurodiverse needs is awaited. All other children were achieving normal milestones at most recent follow-up (median 3 years old, range 0.5-5).

## Discussion

Overall, we have found a relatively small and still developing literature. The evidence was of sufficient quality to synthesise, but compared to our idealised checklist, there was considerable missing data. While our conclusions are largely reassuring, the strength of the evidence is low and should be considered provisional. However, given the prevalence of case reports (45/48, 94%) a format that is intrinsically potentially biased towards atypicality and concern, their low frequency offers a degree of reassurance.

Maternal outcomes did not differ significantly from a systematic review of the disease overall, with 36/51 (71%) of pregnancy-associated cases either fully recovering or with minimal deficit versus 918/1284 (72%) with mRS 0-2. The rate of maternal deaths was also similar (4/51, 8%, versus 81/1284, 6%) (20). The NMDAR-Ab-E cases with onset *before* pregnancy were generally less severe. This could plausibly reflect a selection bias of sufficient recovery to allow subsequent pregnancy. With regard to pregnancy outcome in cases with an onset *during* pregnancy, three miscarriages occurred during the first trimester, and one in the second trimester, broadly in keeping with spontaneous fetal loss (15% in the first trimester and 1-2% in the second) (24,25). While the stillbirth rate was higher than background (5.7% (2/35) vs 0.4%; P = 0.0171, binomial test) (26), this is lower than has been reported for pregnant women admitted to intensive care (9.9%) (27). Moreover, the small sample size of this rare sub-group of a rare disease influenced by reporting bias considerably caveats this comparison.

It is encouraging that the literature and our own experience find cases of pregnancy following resection of ovarian teratoma to treat acute NMDAR-Ab-E. Despite published reports of ovary-preserving surgery (28), given the risk of residual teratoma tissue driving ongoing disease or relapse, oophorectomy remains common. Therefore, consideration of preserving oocytes in young women who have yet to start a family is important. Our multi-disciplinary approach includes a specialist gynaecologist with expertise in both teratoma resection and ovarian cryo-preservation to discuss options with patients and their next of kin (29).

Despite the evidence from animal models, in the available published data we found little positive evidence of developmental disorders in children born to mothers in any of the three sub-groups. We found one reported case of developmental delay reported in 53 (2%) livebirths, broadly in keeping with the frequency amongst children under five in the general population (1-3%) (30). Moreover, whilst this infant’s serum was positive for NMDAR-IgG, the infant was also born prematurely secondary to uteroplacental insufficiency and the mother had severe illness and died of secondary infection (12). Thus, the potential specific effects of transplacental transfer of NMDAR-IgG in this case are challenging to disentangle from other relevant factors. Similarly, in our case series of children born to mothers with either active or previous NMDAR-Ab-E, only one is being evaluated for neurodiverse needs and there was no detectable maternal seropositivity during the pregnancy.

Conversely, the quality of the data we have identified makes it impossible to fully exclude an increased risk of neurodevelopmental conditions. Firstly, specific neonatal health outcomes are often missing from reports (Fig. 4C). Secondly, the data on long term outcomes are very limited and human neurodevelopment manifests over years-decades. Moreover, outside of our re-contacts data, duration of follow-up has not been updated in the literature. Additionally, the type of cohort study design needed to truly determine the effect of autoantibody transfer to be adequately powered and control sufficiently for confounding variables would require multi-centre co-ordination. Pregnancies occurring after full disease remission may be amenable to this but those complicated by NMDAR-Ab-E during or after are by definition heavily confounded by the effects of the disease on maternal and placental condition as well as by the multiple supportive medical, interventional, and immunotherapeutic interventions needed to survive and recover from the illness.

Finally, testing of trans-placental autoantibody transfer remains rare. Compromised neonates were disproportionately likely to be tested, making up five of ten (50%) cases tested despite only seven of the 53 (13%) cases reporting a compromised infant. Four of the five comprised infants tested were positive for NMDAR-IgG, but two went on to meet their developmental milestones at one year of age, whilst one had global developmental delay, and the other died during the neonatal period (Supplementary Table 5). Both the latter had relatively high titres at 1:320 and 1:450 respectively, substantially higher than the 1:20 titre of one of the infants who developed normally but comparable to the other where the titre was 1:400. Furthermore, one healthy infant tested positive for serum NMDAR-IgG (31). Autoantibody testing was available for one neonate in our cohort, and whilst compromised at birth they too have subsequently developed normally. Thus, the clinic-pathologic sequelae of transplacental NMDAR-IgG in humans remains to be fully elucidated. Certainly, NMDARs are important in the developing fetal brain (32). Evidence from mouse models indicates that transplacental transfer of patient derived NMDAR-IgG can result in reduced survival rates in the postnatal period, with reduced brain volume and neurodevelopmental abnormalities in adulthood (5). However, human and murine neurodevelopment differ with an established blood-brain barrier forming postnatally in mice and between 22 and 32 weeks’ gestation in humans (33).

Overall, the reliance on reports introduces a reporting bias and unsystematic reporting. The development of a global, confidential registry to systematically record disease presentation, treatments and disease course, in addition to obstetric, maternal, neonatal and childhood outcomes could improve evidence quality. This has precedent in obstetric practice with caesarean scar pregnancies and within neurology for multiple sclerosis (34,35). Our idealised checklist could serve as a starting point, with further input from existing international clinical-research networks including multi-disciplinary expertise supported by patient advocacy organisations.

## Authors contributions

Conceptualisation – SLH, DS, HF, AAD

Data collection and curation – SLH, SNMB, PC-W, AAD

Formal analysis – SLH, AH, AAD

Writing - original draft – SLH, AAD

Writing - review & editing – All

## Data availability statement

Further data supporting the study are available from the corresponding authors upon reasonable request.

