## Supplementary material for "A systematic review of clinical data and reporting quality in NMDAR-antibody encephalitis and pregnancy"

Supplementary_Table_1.xlsx

**Supplementary Table 1:** Extracted data from published record according to idealised reporting checklist. Data frame of extracted data from included records is given in the first tab. Each pregnancy is reported on a separate row. Excluded full text articles are listed in the second tab.

| **Reason For Exclusion** | **Search date** | | |
| --- | --- | --- | --- |
|  | **21/3/23** | **18/7/23** | **19/10/23** |
| Basic Science/Animal Study | 23 |  | 1 |
| Book Chapter | 8 |  |  |
| Comment piece/Editorial | 33 | 1 | 1 |
| Non-pregnant/male cases | 66 | 1 | 3 |
| Not in English | 13 |  |  |
| Other | 6 |  |  |
| Paediatric Case/Study | 472 | 26 | 13 |
| Review Article | 160 | 8 | 3 |
| Study – Comparison Paediatric and Adult Cases | 9 |  |  |
| Study – Cerebrospinal fluid contents | 27 |  | 1 |
| Study – Epidemiological | 6 |  |  |
| Study – Genetics | 2 |  |  |
| Study – Imaging | 16 |  | 3 |
| Study – Laboratory Diagnosis | 13 |  |  |
| Study – Presenting/Clinical Features | 42 |  | 1 |
| Study – Scoring Systems | 6 |  | 1 |
| Study – Serum | 11 | 1 | 1 |
| Study – Treatment | 11 |  | 2 |
| Wrong disease | 4 | 2 | 1 |

**Supplementary Table 2:** Reason for exclusion of records in initial and subsequent searches – number of records stated under date of corresponding search.

| **Study** | **Case Type** | **Outcome** | **Details** |
| --- | --- | --- | --- |
| Chan et al. | During | Miscarriage | Presented in first trimester, miscarried within two days of hospitalisation. |
| Pennington et al. | During | Miscarriage | Miscarriage in first trimester whilst in early stages of disease. |
| Kim et al. | During | Miscarriage | Miscarried whilst in ICU, cause unclear. |
| Zhang et al. | During | Miscarriage | Miscarriage due to uterine haemorrhage at 16 weeks. Second presentation of NMDAR-Ab-E in pregnancy. |
| Reisz et al. | During | Stillbirth | Stillbirth at 27 weeks, in context of sepsis with pyothorax secondary to central venous catheter. |
| Keskin et al. | During | Stillbirth | Vaginal bleeding and hypotension at 32 weeks. US showed fetal demise and mother subsequently died from sepsis. |
| Kumar et al. | During | Termination | Termination whilst intubated, patient unable to consent. |
| Liu et al. | During | Termination | NMDAR-Ab-E during third and fourth pregnancy. Termination at 15 weeks both times during acute illness when patient unable to consent. Husband consented in first pregnancy. |
| Kittichanteera et al. | During | Termination | Unexpected first trimester pregnancy discovered. Family gave consent for termination, patient acutely unwell and unable to consent. |
| Kokubun et al. | During | Termination | Patient in coma, family gave consent for termination. |
| Tian Nie et al. | During | Termination | Patient recovered sufficiently to consent to termination. |
| Wallengren et al. | Before | Termination | Accidental pregnancy after recovery, patient decision for termination. |

**Supplementary Table 3:** Circumstances of miscarriage, stillbirth, and terminations identified within reported literature

Abbreviations: ICU=intensive care unit, NMDAR-Ab-E=NMDAR-antibody encephalitis, US=ultrasound

| **Study** | **Case Type** | **Details** |
| --- | --- | --- |
| Jagota et al. | Intrapartum | Sepsis secondary to aspiration pneumonia. Treated with methylprednisolone and IVIG. Immunotherapy limited by infection. |
| Keskin et al. | Intrapartum | Sepsis secondary to fetal demise. Treated with methylprednisolone and plasmapheresis. Immunotherapy limited by infection. |
| Kittichanteera et al. | Intrapartum | Sepsis after initial recovery following treatment with IVIG and methylprednisolone. |
| Chen et al. | Postpartum | Multi-organ failure in context of severe illness not responsive to IVIG, methylprednisolone, rituximab, plasmapheresis and removal of ovarian teratoma. |

**Supplementary Table 4:** Circumstances of maternal death identified within reported literature

Abbreviations: IVIG=intravenous immunoglobulin

| **Report** | **Illness onset relative to pregnancy** | **Neonatal Outcome** | **NMDAR-IgG transfer tested?** | **Childhood Outcome** |
| --- | --- | --- | --- | --- |
| Lamale-Smith et al. | During | Respiratory and neuromuscular depression likely secondary to maternal drugs, SVT requiring digoxin. | Yes.  Fetal cord blood positive (1:20). | “Doing well and developmentally appropriate” |
| Jagota et al. | During | Intermittent episodes of spontaneous fine movements controlled by phenobarbital. | Yes.  Neonatal serum positive two days after birth, (1:450). | "Delayed in global development and experienced generalized seizures" |
| Kumar et al. | During | Low AGPAR score, 3 at 1 minute, 6 at 5 minutes. | Yes.  Not detected in neonatal serum, CSF or cord blood. | "Met all developmental milestones to date" |
| Scorrano et al. | During | Respiratory distress, hypoglycaemia. Brain ultrasound imaging showed periventricular hyper-echogenicity and sacral ultrasound imaging documented spina bifida. CXR showed right middle and lower lobe and left lower lobe consolidation. | Not Stated | Childhood outcome not stated. |
| Joubert et al. | During | Respiratory insufficiency recovering spontaneously at 24 hours, presumed secondary to maternal drugs. | Not Stated | "Normal behaviour and development" |
| Chourasia et al. | Before | hypotonia, poor respiratory efforts requiring intubation and ventilation, diffuse encephalopathy on EEG. MRI showed diffuse cerebral oedema and extensive ischaemic and haemorrhagic injury. Given IVIG for five days. | Yes.  Neonatal serum positive (1:320). | Neonatal death |
| Hilderink et al. | Before | Respiratory insufficiency. Moro, rooting, grasp, suck and swallow reflexes decreased but muscle tone normal. Required oxygen and gastric tube feeding. IV anti-biotics for 3 days. Opened eyes at day 6 and began feeding, feeding problems resolved by day 10. | Yes.  Neonatal serum positive three days after birth (~1:400). | "At 12 months development was normal" |

**Supplementary Table 5:** Details and outcomes of compromised neonates identified within reported literature. NMDAR-IgG assay end-point dilution stated in the report is noted.

Abbreviations: APGAR=appearance, pulse, grimace, activity and respiration, CXR=chest x-ray, CSF=cerebrospinal fluid, EEG=electroencephalogram, IVIG=intravenous immunoglobulin, NMDAR-IgG=Immunoglobulin G autoantibody against N-Methyl D-Aspartate receptor, SVT=supraventricular tachycardia

| **Case** | **Disease onset relative to pregnancy** | **Age range at NMDAR-Ab-E onset (years)** | **Ovarian teratoma?** | **Age range at pregnancy onset** | **Conception** | **Trimester at NMDAR-Ab-E onset** | **Total admission duration range (months)** | **Delivery time days post-NMDAR-Ab-E onset (months)** | **Pregnancy outcome** | **Mode of delivery** | **Gestational age (Term or Preterm)** |
| --- | --- | --- | --- | --- | --- | --- | --- | --- | --- | --- | --- |
| Case 1  Pregnancy 1 | During | 31-35 | Yes | 31-35 | Spontaneous | 2nd | <6 | <1 | Livebirth | EMCS | Preterm |
| Case 2  Pregnancy 1 | Before | 31-35 | Yes | 31-35 | Spontaneous | N/A | <2 | N/A | Livebirth | Spontaneous vaginal delivery | Term |
| Case 2  Pregnancy 2 | Before | As above | Yes | 36-40 | Spontaneous | N/A | As above | N/A | Livebirth | EMCS | Term |
| Case 3  Pregnancy 1 | Before | 21-25 | Yes | 26-30 | Spontaneous | N/A | <1 | N/A | Miscarriage | N/A | N/A |
| Case 3  Pregnancy 2 | Before | As above | As above | 26-30 | Spontaneous | N/A | As above | N/A | Livebirth | Spontaneous vaginal delivery | Preterm |
| Case 3  Pregnancy 3 | Before | As above | As above | 26-30 | Spontaneous | N/A | As above | N/A | Livebirth | Spontaneous vaginal delivery | Term |
| **Supplementary Table 6: Maternal demographics, NMDAR-Ab-E presentation, pregnancy timings and key outcomes.** Abbreviations: EMCS = emergency C-section, GxPx=gravidity and parity, N/A=not applicable, NMDAR-Ab-E=NMDAR-antibody encephalitis | | | | | | | | | | | |

| **Case** | **Initial MRI brain abnormal?** | **Initial EEG abnormal?** | **Initial CSF protein raised?** | **Initial CSF cell count raised?** | **Serum NMDAR-IgG**  **(initial)** | **CSF NMDAR-IgG (initial)** | **Ovarian teratoma identified?** | **ICU admission** | **Immuno-therapy and/or**  **oophorectomy during illness?** | **If RTX, was it pre- or post-delivery?** | **Symptomatic treatments** | **Maternal outcome** | **Length of maternal follow-up since most recent illness episode (years)** |
| --- | --- | --- | --- | --- | --- | --- | --- | --- | --- | --- | --- | --- | --- |
| Case 1  Pregnancy 1 | Yes | Yes | No | Yes | Positive | Positive | Yes | Yes | Immunotherapy  Oophorectomy | Post-delivery | Anti-seizure medication  Anti-psychotic medication | Outpatient neuro-rehabilitation | 1 |
| Case 2  Pregnancy 1 | No | Yes | No | Yes | Positive | Positive | Yes | Yes | Immunotherapy  Oophorectomy | NA | Anti-seizure medication  Anti-psychotic medication | Recovered, no deficit | 7 |
| Case 2  Pregnancy 2 | NA | NA | NA | NA | NA | NA | NA | NA | As above | NA | NA | Recovered, no deficit | As above |
| Case 3  Pregnancy 1 | Yes | N/A | N/A | N/A | Positive | Positive | Yes | No | Immunotherapy  Oophorectomy | N/A | Anti-seizure medication  Anti-psychotic medication | Recovered, no deficit | 7 |
| Case 3  Pregnancy 2 | As above | As above | As above | As above | Seronegative during pregnancy | As above | As above | As above | As above | As above | As above | As above | As above |
| Case 3  Pregnancy 3 | As above | As above | As above | As above | Seronegative during pregnancy | As above | As above | As above | As above | As above | As above | As above | As above |
| **Supplementary Table 6: NMDAR-Ab-E investigations, treatment and maternal outcomes.** Abbreviations: CBA=cell-based assay, CSF=cerebrospinal fluid, CT=computed tomography, EEG=electroencephalogram, MRI= magnetic resonance imaging, N/A=not applicable, NMDAR-IgG=Immunoglobulin G autoantibody against N-Methyl D-Aspartate receptor, PLEX=plasma exchange, RTX=rituximab, US=ultrasound | | | | | | | | | | | | | |

| **Case** | **Fetal growth restriction** | **Hypertensive disorder of pregnancy** | **Gestational diabetes** | **Ante-partum haemorrhage** | **Low-lying placenta** | **Ovarian cyst accident** | **Emergency surgery during pregnancy** | **Preterm or Term birth** | **Cord prolapse** | **Placental abruption** | **Meconium aspiration** | **Chorio-amnionitis** | **Post-partum haemorrhage (>500ml)** | **Blood transfusion** | **Delivery under GA** |
| --- | --- | --- | --- | --- | --- | --- | --- | --- | --- | --- | --- | --- | --- | --- | --- |
| Case 1  Pregnancy 1 | No | Yes | Yes | No | No | No | No | Preterm | No | No | No | No | No | No | Yes |
| Case 2  Pregnancy 1 | No | No | Yes | No | No | No | No | Term | No | No | No | No | Yes | No | No |
| Case 2  Pregnancy 2 | No | No | Yes | No | No | No | No | Term | Yes | No | No | No | Yes, | No | No |
| Case 3  Pregnancy 1 | N/A | N/A | N/A | N/A | N/A | N/A | N/A | N/A | N/A | N/A | N/A | N/A | N/A | N/A | N/A |
| Case 3  Pregnancy 2 | No | No | No | No | No | No | No | Preterm | No | No | No | No | No | No | No |
| Case 3  Pregnancy 3 | No | No | No | No | No | No | No | Term | No | No | No | No | No | No | No |
| **Supplementary Table 6: Antenatal and delivery outcomes.** Abbreviations: GA=general anaesthetic, N/A=not applicable, NMDAR-Ab-E=NMDAR-antibody encephalitis, PPROM=preterm premature rupture of membranes. | | | | | | | | | | | | | | | |

| **Case** | **Testing of placenta or neonate for NMDAR-IgG** | **Neonatal condition** | **Birth weight (nearest kg)** | **Normal APGARs by 10 minutes?** | **Normal cord gas?** | **HIE?** | **SCBU admission** | **Cooling** | **Infection** | **Seizures** | **Breast feeding before discharge** | **Childhood condition – general statement** | **Milestones – on target or delayed** | **Medical diagnoses** | **Neurologic diagnoses** | **ASD or ADHD** | **Any behaviour condition** | **Length of child follow-up (years)** |
| --- | --- | --- | --- | --- | --- | --- | --- | --- | --- | --- | --- | --- | --- | --- | --- | --- | --- | --- |
| Case 1  Pregnancy 1 | No | Apnoeic episodes  Bradycardia  Jaundice  Ascites | 2 | Unavailable | Unavailable | No | Yes | No | No | No | Yes | Good | On target | No | No | No | No | 1 |
| Case 2  Pregnancy 1 | Both detectable on live CBA | TTN | 4 | Yes | Unavailable | No | No | No | No | No | No | Good | On target | No | No | No | No | 5 |
| Case 2  Pregnancy 2 | No | Good | 4 | Yes | Normal | No | No | No | No | No | Yes | Good | On target | Eczema | No | No | No | 1.5 |
| Case 3  Pregnancy 1 | No | N/A | N/A | N/A | N/A | N/A | N/A | N/A | N/A | N/A | N/A | N/A | N/A | N/A | N/A | N/A | N/A | N/A |
| Case 3  Pregnancy 2 | No | Low APGARs  Jaundice  Sepsis | 3 | Yes | Arterial - Abnormal | No | Yes | No | Yes | No | Yes | Good | On target | No | None | Under review | No | 4 |
| Case 3  Pregnancy 3 | No | Good | 3 | Yes | Unavailable | No | No | No | No | No | Yes | Good | On target | No | No | No | No | 2 |
| **Supplementary Table 6: neonatal and childhood outcomes.** Abbreviations: APGAR=appearance, pulse, grimace, activity and respiration, ASD=autistic spectrum disorder, ADHD=attention deficit hyperactivity disorder, BE=base excess, CBA=cell-based assay, CRP=C-reactive protein, HIE=hypoxic ischaemic encephalopathy, IV=intra-venous, N/A=not applicable, NMDAR-IgG=Immunoglobulin G autoantibody against N-Methyl D-Aspartate receptor, PN=parenteral nutrition, PPROM=preterm premature rupture of membranes, SCBU=special care baby unit, TTN=transient tachypnoea of the newborn | | | | | | | | | | | | | | | | | | |

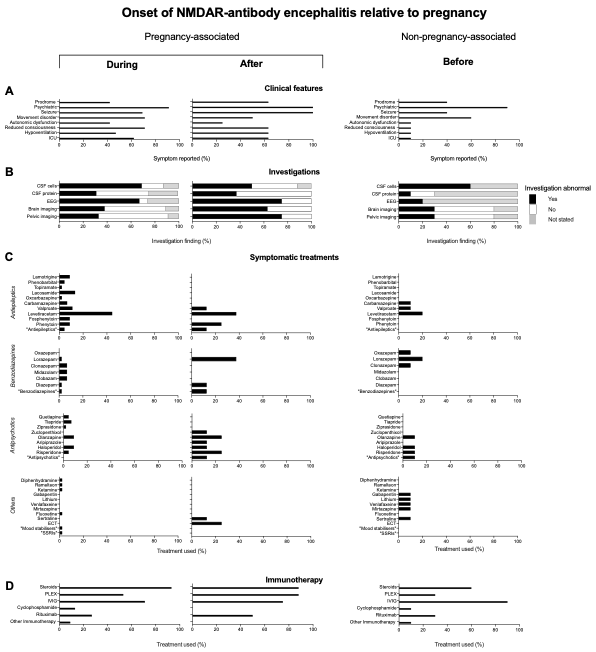

**Supplementary Figure 1: NMDAR-Ab-E clinical features and treatments according to pregnancy association**

A – Rates of clinical features in each group indicated by percentage. B – Rates of investigations and outcomes (abnormal=black, not abnormal=weight; uncertain if investigation done=grey). Brain imaging includes CT or MRI and pelvic imaging includes CT, US, or MRI. C – Rate of symptomatic treatments into anti-epileptics, benzodiazepines, anti-psychotics and other. D – Rates of immunotherapy. Other immunotherapy: during – azathioprine (2), and tacrolimus (1) and bortezomib (1); before – azathioprine (1).

Abbreviations: CT=computed tomography, CSF=cerebrospinal fluid, ECT=electroconvulsive therapy, EEG=electroencephalogram, ICU=intensive care unit, IVIG=intravenous immunoglobulin, MRI=magnetic resonance imaging, PLEX=plasma exchange, SSRI=selective serotonin reuptake inhibitor, US=ultrasound.

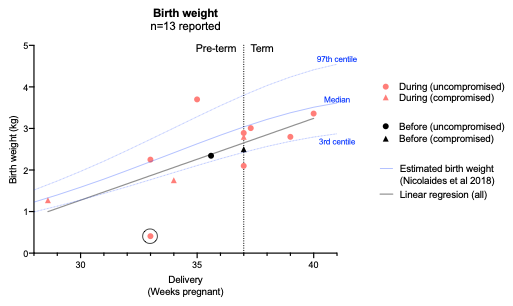

**Supplementary Figure 2: Neonatal birthweights including outlying value**

The plot from Fig. 3C is reproduced to include and analyse an outlying reported value of 408g at 33 weeks (circled) (21). This weight is extremely low for gestational week 33 and premature babies born this small would be unlikely to survive. Attempts to contact authors to clarify birthweight were unsuccessful. The line of best fit consequently moves towards the lower limit of normative values.

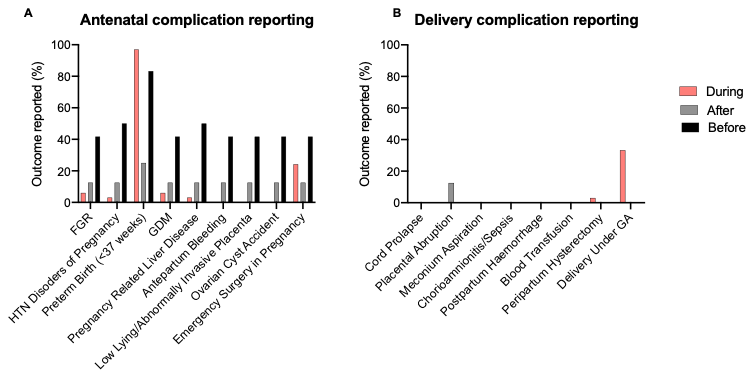

**Supplementary Figure 3: Antenatal and delivery outcome reporting**

A – Percentage of each case type reporting on each of the listed antenatal complications, either positively or negatively. B – Percentage of each case type reporting on each of the listed delivery complications, either positively or negatively.

Abbreviations: FGR=fetal growth restriction, GA=general anaesthetic, GDM=gestational diabetes mellitus, HTN=hypertensive

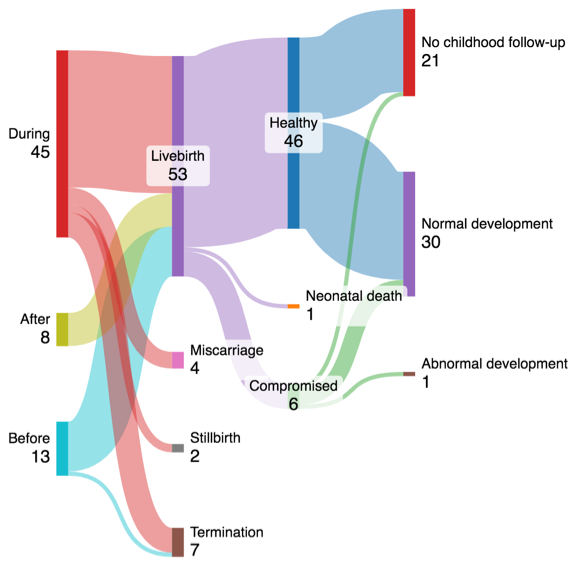

**Supplementary Figure 4: Outcomes summary**

Sankey diagram showing relationship between pregnancy, neonatal and childhood outcomes in mothers who had NMDAR-antibody encephalitis before, during, or after a pregnancy. Numbers of pregnancies are given then after birth numbers relate to offspring.
